# The Imperative for Innovation: Gradient Boosting Capabilities in Diagnosing Ischemic Heart Disease Using Exhaled Breath Analysis

**DOI:** 10.1101/2025.11.01.25339309

**Authors:** Basheer Abdullah Marzoog, Anastasia Stroeva, Philipp Kopylov

## Abstract

**Background:** Ischemic heart disease and the related sequalae remains the leading cause of mortality and morbidity globally. The poor diagnostic accuracy and availability of early screening methods are the leading objectives.

**Aims:** To assess the diagnostic capabilities of the machine leaning model in the diagnosis of ischemic heart disease using the exhaled breath analysis predictors.

**Materials and methods:** A single center prospective study involved participants with vs without stress induced myocardial perfusion defect. All the participants underwent real-time breath analysis using a PTR-TOF-MS-1000, bicycle ergometry test, and multidetector computed tomography (MDCT) of the coronary arteries with myocardial perfusion assessment. The obtained exhaled breath analysis data were analyzed using machine learning model. For statistical analysis used programme Statistica, SPSS, and python.

**Results:** An 80 participants divided into 31 with positive stress-induced myocardial perfusion defect vs 49 without. The diagnostic features of the built model in compare to the reference the MDCT, AUC 86 % (95% confidence interval, [0.7805-0.9338]), sensitivity 0.6129 (95 % CI, [0.4414-0.7844] ), and specificity 0.8367 (95 % CI [0.7332-0.9402]).

**Conclusion:** The Gradient Boosting model shows fascinating results using the exhaled breath analysis in the diagnosis of ischemic heart disease. However, further investigations on a larger sample size are required to uncover the hidden part of the plot.

## Introduction

Ischemic heart disease (IHD), also commonly referred to as coronary artery disease (CAD), remains the preeminent global cause of morbidity and mortality, casting a long shadow over cardiovascular health worldwide [1]. Defined fundamentally as a condition characterized by an imbalance between myocardial oxygen supply and demand, IHD manifests along a spectrum from transient, reversible myocardial ischemia (angina pectoris) to irreversible myocardial infarction (MI) and heart failure [2]. This imbalance is overwhelmingly caused by atherosclerotic plaque formation, progression, and complications (rupture, erosion, thrombosis) within the epicardial coronary arteries, leading to critical luminal narrowing or occlusion [3]. Despite decades of research and clinical advancement, the timely and accurate diagnosis of IHD, particularly in its early or atypical presentations, continues to pose significant challenges, driving an urgent need for transformative diagnostic approaches [4].

The epidemiology of IHD underscores its devastating societal impact. According to the Global Burden of Disease Study, IHD persists as the single leading cause of death globally, responsible for approximately 9 million deaths annually [5]. Its prevalence is staggering, affecting hundreds of millions of individuals. While age-standardized mortality rates have declined in high-income countries due to improved prevention, acute management, and revascularization techniques, the absolute burden continues to rise due to aging populations and increasing prevalence of risk factors like diabetes, hypertension, dyslipidemia, obesity, and sedentary lifestyles, particularly in low- and middle-income countries [6]. Furthermore, a significant proportion of initial presentations are catastrophic events like acute MI or sudden cardiac death, highlighting the critical failure of early detection in current paradigms [7].

The pathophysiology of IHD is centered on coronary atherosclerosis, a chronic inflammatory process initiated by endothelial dysfunction and lipid accumulation within the arterial intima. Progressive plaque growth narrows the vessel lumen, restricting blood flow [8]. Crucially, acute coronary syndromes often arise not from the most severely stenotic lesions, but from plaques deemed “vulnerable” – characterized by a thin fibrous cap, large lipid core, and intense inflammatory activity – which are prone to rupture or erosion, triggering platelet adhesion, aggregation, and thrombus formation, causing sudden, often complete, vessel occlusion [9]. This complex interplay of structural stenosis, endothelial dysfunction (impairing vasodilation during increased demand), inflammation, and thrombosis underlies the diverse clinical manifestations.

However, conventional diagnostic tools often fail to capture this dynamic complexity, particularly the functional significance of non-obstructive plaques or the inflammatory milieu preceding acute events [10].

The limitations of current diagnostic modalities are a key driver of IHD’s persistent dominance. While foundational, the resting electrocardiogram (ECG) lacks sensitivity and specificity for ischemia, especially without active symptoms [11]. Exercise stress testing, while valuable, has variable accuracy, is contraindicated in many, and requires significant patient effort and monitoring resources. Non-invasive imaging like stress echocardiography, myocardial perfusion imaging (MPI), and coronary computed tomography angiography (CCTA) offer improved accuracy but come with substantial drawbacks: high cost, limited accessibility (especially in resource-poor settings), exposure to ionizing radiation (MPI, CCTA), the need for sophisticated equipment and specialized personnel, and often, lengthy wait times. Invasive coronary angiography (ICA), the gold standard for defining coronary anatomy, is expensive, carries inherent procedural risks, and primarily identifies significant luminal narrowing, often missing the functional impact of less severe lesions or the underlying plaque vulnerability. These limitations create significant diagnostic gaps, leading to delayed diagnosis, missed opportunities for early intervention, unnecessary procedures, and ultimately, preventable adverse outcomes. Overcoming the predominance of IHD necessitates a paradigm shift towards accessible, non-invasive, cost-effective, and highly sensitive tools capable of detecting the disease in its nascent or subclinical stages, particularly within the high-throughput environment of the outpatient department (OPD) [12].

This imperative has catalyzed the exploration of novel diagnostic frontiers. Among the most promising is exhaled breath analysis (EBA). Human breath contains a complex mixture of volatile organic compounds (VOCs), metabolic end-products reflecting physiological and pathophysiological processes occurring throughout the body, including the heart. Research indicates that specific VOCs, or distinct patterns (breathprints), are generated during myocardial ischemia due to oxidative stress, inflammation, lipid peroxidation, and shifts in energy metabolism [13–15]. The profound appeal of EBA lies in its inherent characteristics: it is completely non-invasive, requiring only a simple breath sample; painless and safe for patients; rapid (results potentially available in minutes); low-cost compared to imaging; and highly accessible, requiring minimal infrastructure beyond the collection device and analyzer. This makes it uniquely suited for large-scale screening programs in the OPD setting [16]. However, interpreting the complex VOC signatures within breath presents a formidable analytical challenge.

This is where the transformative power of machine learning (ML) converges with EBA. ML algorithms, particularly sophisticated pattern recognition techniques like deep learning, possess an unparalleled ability to discern subtle, multidimensional patterns within vast and complex datasets – precisely the nature of breathomics data [17]. By training ML models on breath samples from well-characterized cohorts (confirmed IHD patients vs. healthy controls), these algorithms can learn to identify the unique “breathprint” associated with ischemic heart disease with high sensitivity and specificity. ML can integrate EBA data with other readily available clinical parameters (age, risk factors, basic biomarkers) to create even more powerful predictive and diagnostic models [18]. The potential impact is dramatic: ML-enhanced EBA could revolutionize IHD diagnostics by enabling ultra-early detection of underlying ischemia or vulnerable plaques before significant symptoms or catastrophic events occur. Deployed in the OPD, this technology could facilitate rapid, point-of-care screening, allowing clinicians to efficiently triage patients at high risk for expedited, more targeted investigation (like CCTA or stress testing), while confidently reassuring low-risk individuals [19]. This shift towards proactive, accessible screening holds the potential to significantly reduce the diagnostic gap, mitigate the overwhelming burden of IHD, and usher in a new era of precision cardiovascular medicine focused on true prevention. The convergence of breath analysis and artificial intelligence represents not just an incremental improvement, but a potential revolution in our approach to conquering the world’s leading cause of death [20].

## Material and methods

### Study design

This prospective single-center study enrolled 80 participants: 31 exhibiting stress-induced myocardial perfusion defects and 49 without. Conducted according to Good Clinical Practice (GCP) standards and the Helsinki Declaration, the study received approval from the Sechenov University ethics committee (Protocol No. 19-23, Oct 26, 2023) and was registered on ClinicalTrials.gov (NCT06181799), where inclusion/exclusion criteria are available. All participants provided written informed consent for both the study and personal data processing. Contrast-enhanced CT perfusion imaging results under adenosine triphosphate (ATP) stress were assessed by physicians with at least two years of clinical experience.

The study design involved performing physical exertion test to make us able to assess the dynamic in the volatile organic compounds and calculate delta instead of just at rest assess the VOCs. This makes the study more accurate while physical exertion induced ischemia make us finding the most important VOCs that are associated with ischemic heart disease and not artifacts.

### Data collection

1. Screening – Patient interviews, collection of medical and lifestyle history, review of medical records, physical examination, and documentation of height, weight, blood pressure, pulse, and heart rate.
2. All participants underwent real-time breath analysis at rest using a PTR-TOF-MS-1000 instrument (IONICON PTR-TOF-MS-1000 Trace VOC Analyzer, Austria). Testing occurred in the hospital morning under fasting conditions (6–8 hours), with no toothbrushing permitted [21]. Participants used sterile disposable mouthpieces; no additional filters were required per manufacturer guidelines. Each provided a 1-minute breath sample (yielding 12–16 exhalation cycles). Molecules were ionized using H O primary ions, separated by their mass-to-charge ratio (m/z), and detected. Full-scan mass spectra covered m/z 10–685 (scan time: 1000 ms). The drift tube (T-Drift) and inlet (T-Inlet) temperatures were maintained at 80°C.
3. Study participants passed exercise bicycle ergometry test (on SCHILLER CS 200 device; Bruce protocol) to evaluate the response to physical activity. According to the results metabolic equivalent and Watt, the angina functional class (FC) in participants with positive physical stress test results determined, where watts/Mets <50/<4 FC-III, watts/Mets 50-100/4-7 FC-II, watts /Mets >100/7 FC-I. During bicycle ergometry, the participants monitored with 12-lead electrocardiogram (ECG) and manual blood pressure measurement, one measured at the end of each 2 minute.

The ergometry procedure was stopped if an increase in blood pressure >220 mmHg or horizontal or downward ST segment on the ECG ≥ 1 mm. Furthermore, stop the procedure if the target heart rate (86% of the 220-age) is reached.

1. Prior to undergoing multidetector computed tomography (MDCT) of the coronary arteries with myocardial perfusion assessment, all patients were required to provide laboratory test results indicating serum creatinine levels, followed by calculation of the glomerular filtration rate (GFR) using the Chronic Kidney Disease Epidemiology Collaboration (CKD-EPI) formula. The GFR was required to be no lower than 30 mL/min/1.73 m² to ensure eligibility for the procedure [22–25]. Subsequently, radial vein catheterization was performed to administer a contrast agent and sodium adenosine triphosphate (ATP), with the objective of inducing myocardial ischemia by elevating heart rate.

Multidetector computed tomography (MDCT) of the coronary arteries with myocardial perfusion assessment was performed using a Canon Aquilion One Genesis scanner (Japan; manufacturer: Canon Medical Systems Corporation; device registered with Roszdravnadzor under No. RZN 2021/16161, dated 28 December 2021). The protocol included 640 slices with a native (non-contrast) slice thickness of 0.5 mm, followed by contrast-enhanced imaging to evaluate myocardial perfusion at rest and 20 minutes after intravenous administration of Triphosadeninum (sodium adenosine triphosphate [ATP]; Russia; manufacturer: Ellara; registration certificate No. LP-004667, dated 25 January 2018).

The contrast agent used was Iohexol, 50 mL (Omnipaque; Norway; manufacturer: GE Healthcare; registered with Roszdravnadzor under No. P N015799/01, linked to LP-N(008665)-(RG-RU), dated 14 May 2009). Sodium adenosine triphosphate (3 ampoules, each containing 1 mL [10 mg/mL]) was diluted in 17 mL of 0.9% sodium chloride solution. The resulting 20 mL solution was administered intravenously via slow bolus over 2 minutes at a dose of 300 μg/kg (adjusted for body weight: 60 kg – 12 mL, 70 kg – 14 mL, 80 kg – 16 mL, 100 kg – 20 mL).

Cardiothoracic radiologists performing the CT myocardial stress perfusion analysis were blinded to the results of the stress ECG test.

Image interpretation followed the standardized 16-segment myocardial model endorsed by the American Heart Association, with layered analysis of basal, mid-ventricular, and apical cardiac segments. Initial segmental evaluation was conducted in the short-axis projection to identify artifacts complicating diagnostic interpretation, followed by detailed analysis for perfusion defects [12].

The topographic distribution of coronary blood supply was aligned with the following anatomical landmarks:

- Segments 1, 2, 7, 8, 13, and 14 were assigned to the perfusion territory of the left anterior descending artery (LAD).
- Segments 5, 6, 11, 12, and 16 corresponded to the vascular territory of the circumflex branch (LCx) of the left coronary artery.
- Segments 3, 4, 9, 10, and 15 were associated with the perfusion territory of the right coronary artery (RCA).

Myocardial perfusion analysis involved the visual identification of regions of relative hypoperfusion. Areas of reduced X-ray attenuation, visualized during the arterial phase following adenosine triphosphate (ATP) administration, were interpreted as ischemic changes.

Subsequently, automated software analysis was performed using the Vitrea Advanced platform (Vitrea Workstation). The transmural perfusion ratio (TPR) was calculated, and polar maps were generated to illustrate the distribution of myocardial X-ray attenuation at rest and under stress. Global and segmental perfusion assessments were based on TPR values. A five-color scale was employed to visualize perfusion defect severity:

- Blue (TPR 2.5–0.99): Normal perfusion;
- Green (TPR 0.99–0.97): Minimal perfusion abnormalities;
- Yellow (TPR 0.97–0.94): Moderate hypoperfusion;
- Orange (TPR 0.94–0.60): Significant hypoperfusion;
- Red (TPR 0.60–0.20): Absent perfusion.

Stress perfusion results were considered positive if a stress-induced perfusion defect (TPR <0.97) was observed in one or more myocardial segments.

Multidetector computed tomography (MDCT) with perfusion stress testing served as the reference method for diagnosing coronary artery disease (CAD), in accordance with the 2024 guidelines of the Russian Society of Cardiology (RSC), European Society of Cardiology (ESC), and American College of Cardiology (ACC) [26–28].

Following completion of all study phases, all patients were referred for a follow-up cardiology consultation to adjust or initiate treatment based on the diagnostic findings.

### Statistical analysis

Following data collection, a comprehensive database was compiled. Statistical analysis included descriptive statistics: for quantitative parameters, normality (assessed via Shapiro-Wilk test), mean, standard deviation, median, interquartile range (IQR), minimum, and maximum; for categorical variables, frequencies, and percentages. Comparative analysis used Welch’s t-test for normally distributed data and the Mann-Whitney U-test for non-normally distributed data. Analyses were performed using Statistica 12 (StatSoft, Inc., 2014) and IBM SPSS Statistics (version 29.0.1.1, IBM Corp., 2024), with statistical significance set at p < 0.05. The study design and statistical approach were rigorously aligned with the research objectives and hypotheses.

### Method of Machine learning method building

The Gradient Boosting model demonstrated superior performance in predicting myocardial perfusion defects following adenosine triphosphate stress testing using breath analysis-derived metabolic features, achieving a high AUC through a rigorous methodology that encompassed comprehensive feature engineering, advanced data preprocessing, and robust validation. Delta features (Δ10 and Δ30) were calculated for each metabolite by normalizing post-stress measurements against baseline values, with interaction terms generated between top-variance delta features to capture nonlinear relationships. The preprocessing pipeline excluded features with >50% missing values, applied K-nearest neighbors imputation (k=5) for remaining missing data, addressed class imbalance via SMOTE oversampling exclusively during training folds, and encoded target classes (“Yes”/”No”). Feature selection utilized Recursive Feature Elimination with Gradient Boosting (RFE-GB) to identify the 15 most predictive features from 186 engineered delta features. The optimized Gradient Boosting classifier (n_estimators=200, subsample=0.8, max_depth=5) was evaluated against XGBoost and Random Forest algorithms using Leave-One-Out Cross-Validation (LOOCV) with embedded SMOTE to prevent data leakage, ultimately demonstrating the highest diagnostic with clinically relevant specificity and NPV, while maintaining sensitivity and PPV, with feature importance analysis identifying key metabolic delta features driving predictions. This comprehensive validation framework ensures model generalizability for clinical deployment.

## Results

### Descriptive features of the sample

The study initially enrolled 101 patients. Following the application of predefined exclusion criteria, 21 patients were excluded, yielding a final cohort of 80 participants. Participants were stratified into two groups based on the transmural perfusion ratio (TPR):

Group 1: Stress-induced perfusion defects (TPR ≤ 0.97; n=31);

Group 2: No stress-induced perfusion defects (TPR > 0.97; n=49).

The initial study phase lasted approximately 1.5–2 hours for all participants. The mean waiting time for myocardial perfusion CT was 24.85 days (±23.472 SD). The mean interval between the initial phase (cardiologist consultation with single-channel ECG and plethysmography) and the perfusion CT phase was 21.41 days (±24.09 SD) in Group 1 and 27.02 days (±23.053 SD) in Group 2. This inter-group difference was not statistically significant (*p* = 0.301). Comparative group characteristics are presented in Tables 1 and 2.

**Table 1.** Comparative characteristic of the patients of both groups.

| Index | Whole sample mean ± SD | 1<sup>st</sup> group mean ± SD | 2<sup>nd</sup> group mean ± SD | P |
| --- | --- | --- | --- | --- |
| Age (years) | 56.28 ± 10.60 | 59.93 ± 11.70 | 53.96 ± 9.23 | 0.013* |
| Pre-test probability of IHD (%) | 8.81 ± 7.71 | 9.90 ± 8.14 | 8.12 ± 7.42 | 0.31 |
| HR (beat/min) | 70.29 ± 9.55 | 70.22 ± 10.74 | 70.32 ± 8.84 | 0.96 |
| SBP at rest (mmHg) | 123.16 ± 15.43 | 124.48 ± 20.56 | 122.32 ± 11.22 | 0.54 |
| DBP at rest (mmHg) | 80.61 ± 11.23 | 82.35 ± 13.16 | 79.51 ± 9.81 | 0.27 |
| Weight (kg) | 77.92 ± 16.23 | 77.12 ± 14.71 | 78.42 ± 17.25 | 0.73 |
| height (cm) | 169.95 ± 8.83 | 169.80 ± 9.41 | 170.04 ± 8.54 | 0.90 |
| BMI (kg/m<sup>2</sup>) | 26.93 ± 4.90 | 26.70 ± 4.23 | 27.06 ± 5.31 | 0.75 |
| Target heart rate (bpm) | 163,72 ± 10,60 | 160,06 ± 11,70 | 166,03 ± 9,23 | 0,013* |
| Maximum heart rate (bpm) | 146,25 ± 14,11 | 142,67 ± 17,66 | 148,51 ± 10,91 | 0,071 |
| Percentage of max. HR | 89,53 ± 9,16 | 89,42 ± 12,09 | 89,60 ± 6,84 | 0,93 |
| WT | 125,63 ± 44,11 | 120,96 ± 47,03 | 128,57 ± 42,38 | 0,45 |
| METs | 6,67 ± 1,97 | 6,32 ± 2,03 | 6,88 ± 1,92 | 0,22 |
| Ejection fraction (%) | 64.52 ± 4.38 | 64.47 ± 4.77 | 64.54 ± 4.22 | 0.95 |
| Creatinine (μmol/L) | 82.74 ± 16.01 | 80.37 ± 15.44 | 84.23 ± 16.34 | 0.29 |
| eGFR | 85.31 ± 14.68 | 84.85 ± 15.10 | 85.58 ± 14.53 | 0.82 |
| Statistical significance was defined as p < 0.05<br>Continuous variables are reported as mean ± standard deviation (SD).<br>Independent samples were compared using Student's t-test.<br>Abbreviations: systolic blood pressure (SBP), diastolic blood pressure (DBP), body mass index (BMI), heart rate (HR), estimated glomerular filtration rate (eGFR), ischemic heart disease (IHD), Watt (WT), metabolic equivalent (MET) | | | | |

**Table 2.** A comparative analysis of categorical variables was conducted between patients with stress-induced myocardial perfusion defects (Group 1) and those without such defects (Group 2).

| **Variable** | **Factor** | **Whole sample**<br>**n(%)** | **1st Group**<br>**n(%)** | **2nd Group**<br>**n(%)** | **p** |
| --- | --- | --- | --- | --- | --- |
| Gender | M | 41<br>(51,25%) | 14<br>(45,16%) | 27<br>(55,10%) | 0,38 |
| Pretest probability of IHD | Low | 26 (<br>32,50 %) | 7 (<br>17,07 %) | 19 (<br>30,64 %) | 0,31 |
|  | Moderate | 40 (<br>50,00%) | 18 (<br>43,90 %) | 22<br>(35,48 %) |  |
|  | High | 14 (<br>17,50 %) | 6<br>(14,63 %) | 8 (<br>12,90 %) |  |
| Weight and stage of obesity | Normal | 30<br>(37,50%) | 12<br>(38,70%) | 18<br>(36,70%) | 0,98 |
|  | Overweight | 29<br>(36,25%) | 11<br>(35,48%) | 18<br>(36,73%) |  |
|  | 1st degree | 20<br>(25,00%) | 8<br>(25,80%) | 12<br>(24,48%) |  |
|  | 3rd degree | 1 (1,25%) | 0 (0,00%) | 1 (2,04%) |  |
| Smoking | No | 66<br>(82,50%) | 24<br>(77,41%) | 42<br>(85,71%) | 0,34 |
| Comorbidities | No | 35<br>(43,75%) | 12<br>(38,70%) | 23<br>(46,93%) | 0,83 |
|  | Missing data | 4 (5,00%) | 4<br>(12,903%) | 0 (0,00%) |  |
| Coronary artery atherosclerosis | No | 49<br>(61,25%) | 15<br>(48,38%) | 34<br>(69,38%) | 0,06 |
| Hemodynamically significant coronary artery atherosclerosis by CT with myocardial perfusion assessment (stenosis >70%) | No | 71<br>(88,75%) | 23<br>(74,19%) | 48<br>(97,95%) | 0,002 * |
| Stress-induced myocardial perfusion defect | No | 49<br>(61,25%) | 0 (0,00%) | 49<br>(100,00%) | <0,001 * |
| Defect in myocardial perfusion at rest | No | 54<br>(67,50%) | 10<br>(32,25%) | 44<br>(89,79%) | <0,001* |
| Atherosclerosis of arteries of other localizations | No | 32<br>(40,00%) | 7<br>(22,58%) | 25<br>(51,02%) | 0,006* |
|  | Missing data | 7 (8,75%) | 2 (6,45%) | 5 |  |

| Variable | Factor | Whole sample<br>n(%) | 1st<br>Group<br>n(%) | 2nd<br>Group<br>n(%) | p |
| --- | --- | --- | --- | --- | --- |
|  |  |  |  | (10,20%) |  |
| Arterial hypertension | No | 40<br>(50,00%) | 12 (38,70<br>%) | 28 (57,14<br>%) | 0,10 |
| Stage of arterial<br>hypertension | I | 5 (6,25%) | 4<br>(13,55%) | 1 (2,04%) | 0,07 |
|  | II | 20<br>(25,00%) | 6<br>(19,35%) | 14<br>(28,57%) |  |
|  | III | 16<br>(20,00%) | 9<br>(29,03%) | 7<br>(14,28%) |  |
| Degree of arterial<br>hypertension | Grade 1 | 19<br>(23,75%) | 10<br>(32,25%) | 9<br>(18,36%) | 0,09 |
|  | Grade 2 | 13<br>(16,25%) | 3 (9,67%) | 10<br>(20,40%) |  |
|  | Grade 3 | 9 (11,25%) | 6<br>(19,35%) | 3 (6,12%) |  |
| Antihypertensive drugs | Yes | 23(28,75%) | 12<br>(38,70%) | 11 (22,44<br>%) | 0,11 |
|  | No | 57(71,25%) | 19<br>(61,29%) | 38<br>(77,55%) |  |
| Angiotensin II receptor<br>blockers | Yes | 10 (12,50%) | 6<br>(19,35%) | 4 ( 8,16%) | 0,14 |
|  | No | 70 (87,50<br>%) | 25 (80,65<br>%) | 45 (91,83<br>%) |  |
| Angiotensin-converting<br>enzyme inhibitors | Yes | 7 (8,75%) | 3 (%) | 4 ( 8,16%) | 0,81 |
|  | No | 73<br>(91,25%) | 28 (%) | 45 (91,83<br>%) |  |
| Beta-blockers | Yes | 13<br>(16,25%) | 9<br>(29,03%) | 4 ( 8,16%) | 0,013* |
|  | No | 67<br>(83,75%) | 22 (70,97<br>%) | 45 (91,83<br>%) |  |
| Calcium channel blockers | Yes | 5 (6,25% ) | 3 (9,67 %) | 2 (4,08%) | 0,31 |
|  | No | 75<br>(93,75%) | 28 (90,32<br>%) | 47 (95,91<br>%) |  |
| Diuretics | Yes | 5 (6,25% ) | 3<br>(19,35%) | 2 (4,08%) | 0,31 |
|  | No | 75<br>(93,75%) | 28 (80,65<br>%) | 47 (95,91<br>%) |  |
| History of IHD | Yes | 3 (3,75%) | 2(6,45%) | 1 (2,04%) | <0,001<br>* |
|  | No | 29<br>(36,25%) | 4<br>(12,90%) | 25<br>(51,02%) |  |
|  | Missing data | 48<br>(60,00%) | 25<br>(80,64%) | 23<br>(46,93%) |  |

| Variable | Factor | Whole sample n(%) | 1st Group n(%) | 2nd Group n(%) | p |
| --- | --- | --- | --- | --- | --- |
| Type of Blood Pressure Response to Stress Test | Asthenic | 4 (5,00%) | 2 (10,50%) | 2 (4,08%) | 0,07 |
|  | Hypotonic | 4 (5,00%) | 3 (15,80%) | 1 (2,04%) |  |
|  | Hypertensive | 8 (10,00%) | 1 (5,30%) | 4 (8,16%) |  |
|  | Normotonic | 64 (80,00%) | 1 (5,30%) | 0 (0,00%) |  |
| Watt Functional Class | FCI | 8 (10,00%) | 1 (3,22%) | 7 (14,28%) | 0,31 |
|  | FCII | 9 (11,25%) | 3 (9,67%) | 6 (12,24%) |  |
|  | No IHD | 63 (78,75%) | 27 (87,09%) | 36 (73,46%) |  |
| Functional class according to MET | FCI | 6 (7,50%) | 1 (3,22%) | 5 (10,20%) | 0,55 |
|  | FCII | 10 (12,50%) | 3 (9,67%) | 7 (14,28%) |  |
|  | FCIII | 1 (1,25%) | 0 (0,00%) | 1 (2,04%) |  |
|  | No IHD | 63 (78,75%) | 27 (87,09%) | 36 (73,46%) |  |
| Type of Stress Test Response | Negative | 42 (52,50%) | 16 (51,61%) | 26 (53,06%) | 0,19 |
|  | Suspicious | 21 (22,5%) | 11 (35,45%) | 10 (20,40%) |  |
|  | Positive | 17 (21,25%) | 4 (12,90%) | 13 (26,53%) |  |
| Reason for stopping the stress test | Horizontal ST depression >1 mm | 8 (10%) | 2 (6,45%) | 6 (12,24%) | 0,38 |
|  | Achieving the target heart rate | 72 (90%) | 29 (93,54%) | 42 (88,11%) |  |
| Exercise tolerance | Low | 2 (2,50%) | 1 (3,22%) | 1 (2,04%) | 0,41 |
|  | Moderate | 43 (53,75%) | 20 (64,45%) | 23 (46,93%) |  |
|  | Close to high | 8 (10,00%) | 3 (9,67%) | 5 (10,20%) |  |
|  | High | 16 (20,00%) | 3 (9,67%) | 13 (26,53%) |  |
|  | Very high | 11 (13,75%) | 4 (12,90%) | 7 (14,28%) |  |
| CKD stage | I | 35 (43,75%) | 12 (38,70%) | 23 (46,93%) | 0,28 |
|  | II | 41 | 16 | 25 |  |

| Variable | Factor | Whole sample<br>n(%) | 1st Group<br>n(%) | 2nd Group<br>n(%) | p |
| --- | --- | --- | --- | --- | --- |
|  |  | (51,25%) | (51,61%) | (51,02%) |  |
|  | IIIa | 4 (5,00%) | 3 (9,67%) | 1 (2,04%) |  |
| Lipid-lowering therapy | Yes | 15 (18,75<br>%) | 6 (19,35<br>%) | 9 (14,75<br>%) | 0,91 |
|  | No | 65 (81,25<br>%) | 25 (80,64<br>%) | 40 (65,57<br>%) |  |

\* Statically significant difference values, $p < 0.05$ .
- The categorical variables of the sample are represented as n (%).
- The chi-square test is used as the explanatory variables.
- Abbreviations: MET – metabolic equivalent, CKD – chronic kidney disease, ATP – sodium adenosine triphosphate, HR – heart rate, IHD – ischemic heart disease, FC – functional class

### The Gradient Boosting model performance in diagnosing ischemic heart disease

According to the method that explained in the section “material and method”, the machine learning model was built, and showed a good performance that is clinical acceptable. (Figure 1 and Table 3)

**Figure 1.**
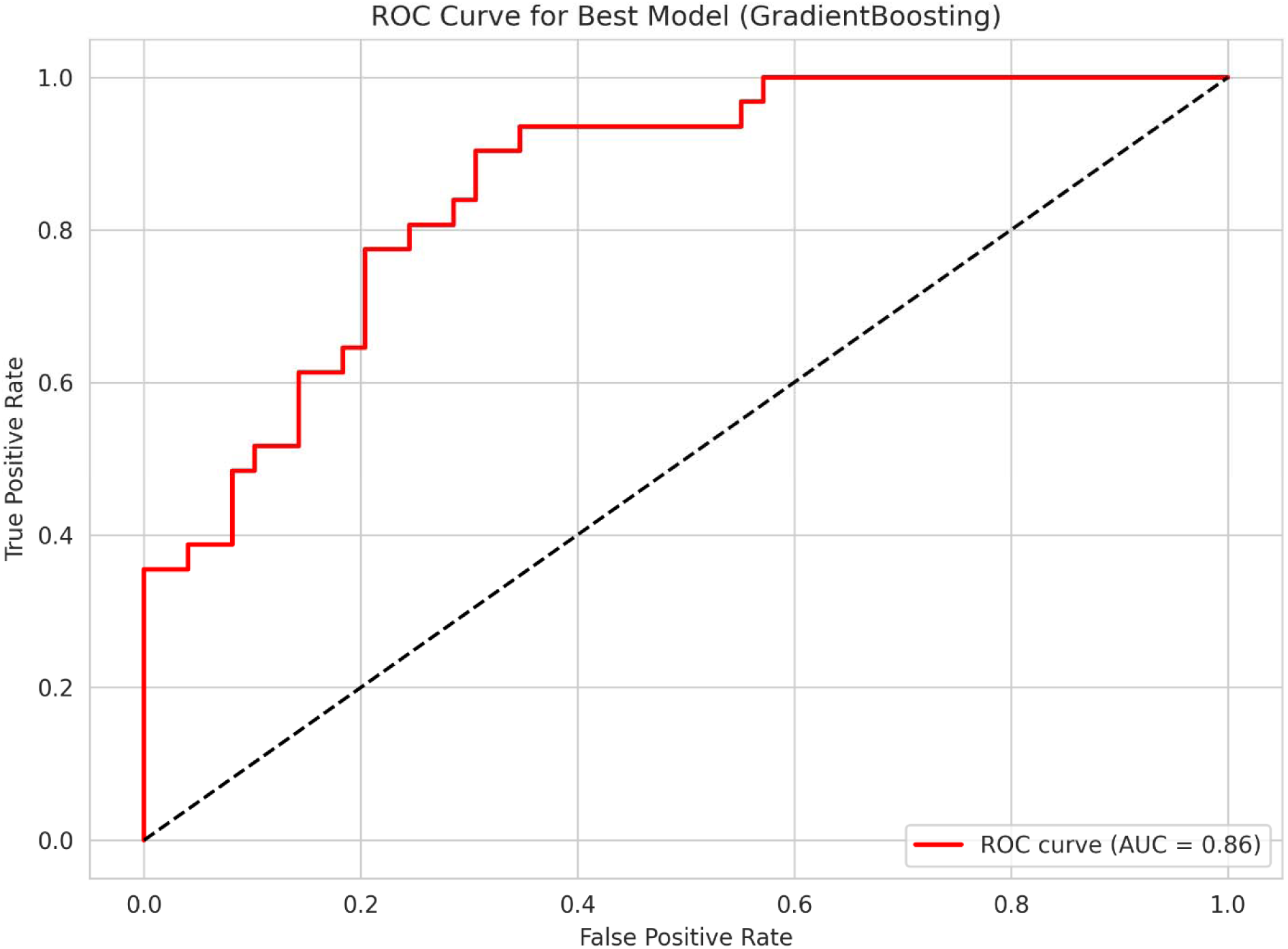
Diagrammatic representation of the built Gradient boosting machine model based on the exhaled breath analysis predictors. The model shows a clinically acceptable results in th diagnosis of ischemic heart disease, AUC 86 %.

**Table 3.** The diagnostic features of the built Gradient boosting machine model based on the exhaled breath analysis predictors.

| Matrices | Mean | 95% Confidence interval |
| --- | --- | --- |
| Accuracy | 0.7500 | [0.6551-0.8449] |
| AUC | 0.8571 | [0.7805-0.9338] |
| Sensitivity | 0.6129 | [0.4414-0.7844] |
| Specificity | 0.8367 | [0.7332-0.9402] |
| PPV | 0.7037 | [0.5315-0.8759] |
| NPV | 0.7736 | [0.6609-0.8863] |

Feature importance, the VOCs that have important role in the diagnosis of ischemic heart disease represented in the table below. (Table 4)

**Table 4.** The feature importance of the obtained volatile organic compounds in the diagnosis of ischemic heart disease based on the built Gradient boosting machine learning model.

| Feature | Coefficient importance |
| --- | --- |
| 27.022364696471204_delta10_x_43.04741898699762_delta10 | 0.116626 |
| 51.03878893892946_delta10 | 0.107111 |
| 132.05190231810434_delta10 | 0.102981 |
| 87.07684847044642_delta10 | 0.102912 |
| 27.022364696471204_delta10_x_47.04077398412186_delta10 | 0.086302 |
| 144.91780932607332_delta10 | 0.071276 |
| 90.95112242970978_delta10 | 0.067960 |
| 144.91780932607332_delta30 | 0.065873 |
| 144.08472746785708_delta10 | 0.053535 |
| 101.94633498924388_delta10 | 0.052952 |
| 118.07092673002178_delta10 | 0.051856 |
| 51.03878893892946_delta30 | 0.049516 |
| 49.99501251131184_delta10 | 0.039997 |
| 37.71047263777778_delta10_x_105.06695512504818_delta30 | 0.021059 |
| 57.06920514918207_delta30 | 0.010044 |

## Discussion

The present study demonstrates the significant diagnostic potential of exhaled breath analysis (EBA) coupled with gradient boosting machine learning (ML) for detecting stress-induced myocardial perfusion defects—a key marker of ischemic heart disease (IHD). Our model achieved an AUC of 0.8571 (95% CI: 0.7805–0.9338) and specificity of 0.8367 (95% CI: 0.7332–0.9402), highlighting its capacity to identify high-risk individuals while minimizing false positives. These findings align with the urgent need for non-invasive, accessible screening tools in IHD diagnostics, particularly given the limitations of conventional methods like stress ECG or CT perfusion imaging, which entail radiation exposure, cost, and accessibility barriers.

Comparing the obtained results with other studies covering the diagnosis of ischemic heart disease using the analysis of the exhaled breath analysis using machine learning model, our model shows the best results. A recent study using the exhaled breath analysis in the diagnosis of ischemic heart disease demonstrated a diagnostic accuracy of 84% [29]. Whereas, another study aimed to assess the existence of coronary artery disease utilizing exhaled breath analysis using a designated portable electronic nose (eNose) system showed a 68% accuracy [30].

## Conclusion

The built Gradient boosting machine learning model based on the exhaled breath data analysis showed a clinically acceptable diagnostic accuracy. However, to confirm the clinical validation of the model, further investigation required to be done to on a larger sample with external validation of the sample.

## Decelerations

1. Ethics approval and consent to participate: the study approved by the Sechenov University, Russia, from “Ethics Committee Requirement № 19-23 from 26.10.2023”. An informed written consent is taken from the study participants.
2. Clinical trial registration: title; Breathome and Single Lead Electrocardiogram Optimizes Ischemic Heart Disease Diagnosis, ID number; NCT06181799, registration date; 2023-12-13 , link to the study; https://clinicaltrials.gov/study/NCT06181799
3. Consent for publication: applicable on reasonable request
4. Availability of data and materials: applicable on reasonable request
5. Competing interests: The authors declare that they have no competing interests regarding publication.

## Authors’ contributions

MB is the writer, researcher, collected and analyzed data, interpreted the results. and revised the final version of the paper, biostatistical analysis of the sample, AS revised the paper, and PhK revised the final version of the manuscript. All authors have read and approved the manuscript.

## Data Availability

All data produced in the present work are contained in the manuscript

## Acknowledgments

not applicable

## Authors’ information

**Basheer Abdullah Marzoog**, World-Class Research Center

«Digital Biodesign and Personalized Healthcare», I.M. Sechenov First Moscow State Medical University (Sechenov University), 119991 Moscow, Russia; postal address: Russia. Moscow, 8-2 Trubetskaya street, 119991, (marzug @mail.ru, +79969602820). ORCID: 0000-0001-5507-2413. Scopus ID: 57486338800. **Anastasia Stroeva,** World-Class Research Center «Digital Biodesign and Personalized Healthcare», I.M. Sechenov First Moscow State Medical University (Sechenov University), 119991 Moscow, Russia; postal address: Russia. Moscow, 8-2 Trubetskaya street, 119991. ORCID: 0009-0002-3694-5295.**Philipp Kopylov,** director of the institute of the Research Center «Digital Biodesign and Personalized Healthcare», World-Class Research Center

«Digital Biodesign and Personalized Healthcare», I.M. Sechenov First Moscow State Medical University (Sechenov University), 119991 Moscow, Russia; postal address: Russia. Moscow, 8-2 Trubetskaya street, 119991. ORCID: 0000-0002-4535-8685. Scopus ID: 6507736224.

The paper has not been submitted elsewhere

Declaration of AI use: not used

## STANDARDS OF REPORTING

STROBE guideline has been followed.

- The TRIPOD+AI standard of reporting for prediction models has been followed

